# Florbetapir PET Measurements of Amyloid Plaque Deposition Using a White Matter Reference Region are More Closely Correlated with Cross-Sectional and Longitudinal Measures of Alzheimer’s Disease-Related Glucose Hypometabolism

**DOI:** 10.1101/2025.09.18.25336126

**Authors:** V Bhargava, M Wang, Y Chen, J Luo, M Weiner, S Landau, W Jagust, Y Su, EM Reiman, K Chen

## Abstract

**Background:** A white matter reference region for in vivo PET-based amyloid imaging may offer advantages due to its larger anatomical size, reduced susceptibility to scatter correction error, and location at the center-of-field view (Lowe et al. 2018, Lopez-Gonzales et al. 2019). The purpose of this study was to compare the association between Florbetapir-measured mean cortical amyloid deposition and an Alzheimer’s disease-related glucose metabolism marker, when two different reference regions are used for Standardized Uptake Value Ratio (SUVR) calculations. We found that mean cortical Florbetapir Standardized Uptake Value Ratios using a white matter reference region (FBP mcSUVR_wm_) were more strongly correlated with the AD-related glucose measure, Hypometabolic Convergence Index (HCI, Chen, et al., 2011) than SUVRs calculated using a cerebellar reference region of interest (FBP mcSUVR_cb_).

**Methods:** Baseline and 2.16±0.37 year follow-up FBP and fluorodeoxyglucose PET data from 1,238 mild AD dementia, mild cognitive impairment (MCI), and cognitively unimpaired (CU) participants from AD Neuroimaging Initiative (ADNI) were used to compare associations between cross-sectional and longitudinal FBP SUVRs and HCI measurements using a whole cerebellar and white matter reference region.

**Results:** Cross-sectionally, partial correlations between mean cortical SUVR and HCI measurements were significantly stronger (Steiger’s Test p<1.0E-16) using a white matter reference region (r=0.59 [p=4.51E-107]) than using a cerebellar reference region (r=0.40 [p=9.30E-46]). Longitudinally, partial correlations between mean cortical SUVR and HCI changes were also significantly stronger (Steiger’s Test p=2.6E-11) using a white matter reference region (HCI/mcSUVR_wm_ r=0.29 [p=1.26E-11]; HCI/mcSUVR_cb_ (r=-0.04 [p=0.36]). Overall, post-hoc within group cross-sectional and longitudinal analysis were significantly stronger when a white matter reference region was used (Cross Sectional Analysis - **CU:** (Steiger’s Test p<0.01); **MCI:** (Steiger’s Test p=4.63E-11); **AD: (**Steiger’s Test p=7.95E-06); Longitudinal Analysis - **MCI: (**Steiger’s Test p=3.94E-04); **AD:** (Steiger’s Test p=0.02)).

**Conclusions:** This study further supports the use of FBP mcSUVR_wm_ measurements in detecting and tracking AD-related amyloid deposition.

## Introduction

Amyloid beta (Aβ) deposition, as acquired by amyloid PET tracers, can be calculated using a semiquantitative cerebral-to-reference region standardized uptake value ratio (SUVR). Notably, the choice of reference region varies between studies. Commonly used reference regions, such as template-based whole-cerebellum, cerebellar gray matter, and pons have traditionally been selected based on the assumption that these regions are devoid of fibrillar Aβ and have similar perfusion properties to target regions-of-interest (*1*). However, limitations of these reference regions include small size and increased influence of scatter or truncation due to proximity of these regions to the edge of the scanner field of view (*1*).

In support of the use of alternative reference regions, we previously demonstrated improved power in detecting longitudinal 18F-Florbetapir measured amyloid beta changes when a subcortical white matter reference region is used compared to cerebellar or pons reference regions(*1*). In a study by Landau et al. in 2015, amyloid beta quantified using white matter as a reference region showed more physiologically plausible changes - as participant’s Aβ status rose their cortical Aβ levels increased – when compared to the cerebellar gray matter or whole cerebellum reference regions (*2*). These studies highlight the importance of reference region selection for amyloid beta semi-quantification, as it can influence precision, accuracy, and ultimately our biological understanding of the role of amyloid beta in AD pathology.

For example, using various reference regions, multiple studies have reported weak or non-significant correlations between glucose hypometabolism and amyloid deposition in AD patients(*3-5*). Hatashita et al. in 2019, found the annual increase in cortical PiB SUVRs was not significantly associated with annual decrease in glucose metabolism in preclinical AD patients(*4*). In 2021, Ehlrich et al. reported no significant associations between glucose metabolism and Aβ deposition in 16 different regions (using F-18 flutemetamol tracer) in AD patients (*5*). Notably, in all these studies, different reference regions were used for amyloid beta SUVR quantification: cerebellum (*3*), (*4*) or a combined cerebellum and pons reference region (*5*).

In this study, we compare the associations between an AD-specific glucose metabolism measure, Hypometabolic Convergence Index (HCI, see our previous report and Methods section below for more details(*7*), and amyloid beta deposition using both cerebellum and white matter as a reference region using an Alzheimer’s Disease Neuroimaging Initiative (ADNI) cohort.

## Methods

### Study Design

Alzheimer’s Disease Neuroimaging Initiative (ADNI) is a longitudinal, multisite study spanning across 57 study sites in the United States and Canada, created to track multiple clinical, genetic, blood-based, CSF and various neuroimaging biomarkers for AD. The ADNI dataset includes elderly control patients, Mild Cognitive Impairment (MCI), and AD patients followed longitudinally. The ADNI dataset used for this study focused on the inclusion of patients who fall under the classical “amnesic” AD category.

### Participants

Our study population included data from total of 1238 participants with the following distribution: between n=379 for CU (n=86 amyloid positive individuals and n=293 amyloid negative individuals), n=592 for MCI, and n=267 for AD, all underwent both FBP and FDG PET scans concurrently. Full inclusion and exclusion criteria can be found at www.adni-info.org. All subjects were between the ages of 55-90 years old, provided their informed consent, and did not have any other neurological condition. MCI patients had a Clinical Dementia Rating (CDR) of 0.5 with a memory box score of at least 0.5 while AD patients had a CDR of 0.5 or 1. The general clinical/behavioral performance of MCI patients was such that an on-site diagnosis of AD could not be made by a physician. NINCDS and ADRDA criteria were followed for AD diagnosis (*6*).

### FBP and FDG PET

Both FDG and FBP PET data were acquired using standardized ADNI protocols on various PET scanners. Data were corrected for radiation-attenuation and scatter using transmission scans or X-ray CT, and reconstructed using reconstruction algorithms specified for each type of scanner as described at www.loni.ucla.edu/ADNI/Data/ADNI_Data.shtml. Acquired images were reviewed, pre-processed and standardized by ADNI PET Coordinating Center investigators at University of Michigan. The images were then uploaded to the Laboratory of Neuroimaging (LONI) ADNI website previously at UCLA and currently at USC, and ultimately downloaded from the LONI website in NIFTI format by investigators at the Banner Alzheimer’s Institute for the analyses in this report.

Details about FBP images can be found at http://adni-info.org. FBP-PET data was acquired in 5-min frames from 50 to 70 min post-injection. FBP PET scans were acquired and pre-processed following the ADNI pipeline (http://adni.loni.usc.edu/methods/pet-analysis-method/pet-analysis/). FBP images were spatially normalized to MNI template using Statistical Parametric Mapping 12 (SPM12).

For FDG-PET, a 30-min dynamic emission scan, consisting of six 5-min frames, was acquired starting 30 min after the intravenous injection of 5 mCi of [18F]FDG. Subjects were instructed to fast for at least 4 hours before the scan and lay quietly in a dimly lit room with eyes open and minimal sensory stimulation. Each center used standardized procedures to identify artifacts and minimize scanner-dependent differences in FDG uptake. During the additional preprocessing stage at Banner Alzheimer’s Institute, automated algorithms were used to register and average each subject’s six 5-min emission frames, transform each subject’s registered image into a 160×160×1.5 mm voxel matrix with sections parallel to a horizontal section through the anterior and posterior commissures (without any adjustment for size or shape), normalize the images for individual variations in absolute image intensity.

### FBP SUVRs and FDG HCIs

SPM12 was used to extract FBP uptake from mean cortical (mcROI) relative to uptake in (1) cerebral white matter regions and (2) cerebellar regions. Masks of these regions are pre-defined in the MNI template in our previous investigation (*1*). Mean Cortical SUVRs (mcSUVRs) were calculated by dividing FBP uptake in mean cortical ROI by FBP uptake in cerebral WM reference region of interest (mcSUVR_wm_) or by FBP uptake in a cerebellum reference region of interest (mcSUVR_cb_). The cerebral white matter reference region included a collection of voxels in the corpus callosum (using the corpus callosum mask from the WFU_PickAtlas toolbox) and centrum semiovale excluding voxels closest to the grey matter or to the ventricles (for more details refer to(*1*)).

For the cerebellar reference ROI, tracer uptake in the whole cerebellum was used.

**For FDG-PET**, instead of using regional measure, we introduced previously and used in this study a global AD-related glucose hypometabolism measure, Hypometabolic Convergence Index (HCI), developed by Chen et al. in 2011 (*7*). HCI was created to reflect the extent to which the pattern and magnitude of a patient’s FDG measured glucose metabolism resembles that of probable AD patients. The HCI computation consists of 3 steps: 1) We first constructed a AD hypometabolism map by comparing FDG PET images of a preselected set of probable AD patients from the ADNI dataset to the FDG PET images of a matched group of healthy controls. Voxel -wise group differences were then transformed into a z-score map to construct an AD hypometabolic map; 2) Next, we constructed individualized z-scored hypometabolic maps for each participant similarly by comparing voxel-wise hypometabolism between the given participant and the same group of healthy controls; 3) Last, we constructed an HCI score for each subject by taking the product of the subjects hypometabolic map and the AD hypometabolic map (voxel-by-voxel multiplication). HCI was then computed as the voxel-wise summation across all voxels where the z-scores from both maps were negative, divided by 10,000. HCI values range from 0 to 15. A higher HCI score (≥ 8.37) indicates that the subject’s FDG-PET glucose metabolism pattern and magnitude is similar to the glucose metabolism pattern and magnitude of a typical AD patient. A lower HCI score (≤ 8.37) indicates that the given subject is less similar to a typical AD patient (*7*).

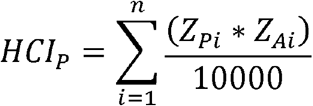

*Z*_*Pi*_ = z score at voxel *i* for person *P, Z*_*Ai*_ is the z score at voxel *i* for AD group and *n* is the total number of voxels associated with hypometabolism in both person of interest and probable AD group within the whole brain volume.

### Statistical Analyses

One-way ANOVA with post-hoc pairwise comparisons was used to evaluate group differences in demographic characteristics including participant scores on the 13-item Alzheimer’s Disease Assessment Scale-Cognitive Subscale (ADAS-Cog-13), a cognitive test assessing multiple cognitive domains including memory, language, praxis, and orientation, and Mini Mental State Exam (MMSE), a brief assessment of mental status assessing orientation, memory, attention, verbal and written ability. A False Discovery Rate (FDR) correction was applied to control for multiple comparisons across variables. Results are summarized in Table 1. We used partial correlation to assess the relationship between global Aβ deposition, using the cerebellum and white matter as reference regions (mcSUVR_cb_ and mcSUVR_wm_), and our AD-related glucose metabolism measure, HCI. Partial correlations were calculated covarying out baseline age and education. A cross-sectional analysis, comparing baseline associations, and a longitudinal analysis, comparing the association between change in HCI and change in mcSUVRs, were each conducted. All observed significant associations remained statistically significant after applying FDR correction.

**Table 1:** Demographics Table.

| Characteristic | CU (n=379) | MCI (n=592) | AD (n=267) | P-Values |
| --- | --- | --- | --- | --- |
| Age (years) | 74 ± 7 | 73 ± 8 | 76 ± 8 | p<0.001 |
| Sex (n [%] female)) | 202 (53%) | 261 (44%) | 113 (42%) | p<0.01 |
| Education (years) | 16.5 ± 2.6 | 16.1 ± 2.7 | 15.9 ± 2.6 | p<0.001 |
| MMSE | 29.0 ± 1.2 | 28.0 ± 1.8 | 22.7 ± 3.4 | p=0.01 |
| ADAS-Cog13 | 9.1 ± 4.6 | 15.3 ± 7.0 | 31.1 ± 9.8 | p<0.001 |
| # of participants with 2 or more visits | 161 | 241 | 101 |  |
| Average number of visits per patient with longitudinal data | 2.05 | 2.22 | 2.17 |  |

Steiger’s Z-test was then used to determine if the difference between partial correlation coefficient of HCI/mcSUVR_cb_ and HCI/mcSUVR_wm_ were significant, with p=0.05. Partial correlations were examined among all subjects and within each diagnostic group: AD, mild cognitive impairment (MCI), and cognitively unimpaired (CU) patients. All statistically significant partial correlations survived an FDR-correction for multiple comparisons.

## Results

Subject characteristics are described in Table 1. The dataset from a total of 1238 participants was included in the cross-sectional analysis of the study with CU (n=379), MCI (n=592), and AD (n=267). All 435 participants with two or more visits, with an average time between visits of 2.16±0.37 years, were included for the longitudinal analysis with CU (n=161), MCI (n=228), and AD (n=46). Patients’ demographic characteristics including sex, education, age, number of months since baseline or visit number, MMSE scores, FBP PET values, and FDG values are shown in Table 1. Significant differences were observed between mean age (p=0.0001), number of females (p=0<0.01), MMSE and ADAS-Cog-13 scores (p=0.01 and p<0.001), and number of years of education (p<0.001 between AD, MCI and CU groups, respectively.

Cross-sectionally, partial correlations between our AD-related glucose measure, HCI, and amyloid deposition were significantly stronger with cerebral white matter reference region than with the cerebellar reference region over all subjects (Figure 1 and Table 2 Overall: HCI/FBP mcSUVR_cb_ r=0.40 (p<0.001) and HCI/FBP mcSUVR_wm_ r=0.59 (p<0.001), Steiger’s test: p<0.001)). Longitudinally, greater partial correlations between HCI/FBP mcSUVR_wm_ were also observed in the entire group (Figure 2 and Table 3 **Overall:** HCI/FBP mcSUVR_cb_ r=-0.04(p=0.36) and HCI/FBP mcSUVR_wm_ r=0.29(p<0.001), Steiger’s Test(p<0.001)).

**Table 2:** Partial Correlations between Cross-Sectional Measurements of Glucose, HCI, and Amyloid Deposition: FBP mcSUVR_wm_ vs FBP mcSUVR_cb_ in CU (n=379), MCI (n=592), AD (n=267), and All Subjects(n=1238).

|  |  | **All Subjects (n=1238)** |  |
| --- | --- | --- | --- |
| **Glucose Measure** | **Amyloid Deposition** | **Partial Correlation** | **Steigers Test** |
| **HCl** | FBP mcSUV<sub>R<sub>cb</sub></sub> | r=0.40 (p=9.30E-46) | p<1.00E-15 |
|  | FBP mcSUV<sub>R<sub>wm</sub></sub> | r=0.59 (p=4.51E-107) |  |
|  | FBP mcSUV<sub>R<sub>cb</sub></sub> and FBP mcSUV<sub>R<sub>wm</sub></sub> | r=0.80 |  |
| **CU (n=379)** |  |  |  |
| **Glucose Measure** | **Amyloid Deposition** | **Partial Correlation** | **Steigers Test** |
| **HCl** | FBP mcSUV<sub>R<sub>cb</sub></sub> | r=0.19 (p=2.97E-04) | p<0.01 |
|  | FBP mcSUV<sub>R<sub>wm</sub></sub> | r=0.40 (p=5.04E-15) |  |
|  | FBP mcSUV<sub>R<sub>cb</sub></sub> and FBP mcSUV<sub>R<sub>wm</sub></sub> | r=0.74 |  |
| **MCI (n=592)** |  |  |  |
| **Glucose Measure** | **Amyloid Deposition** | **Partial Correlation** | **Steigers Test** |
| **HCl** | FBP mcSUV<sub>R<sub>cb</sub></sub> | r=0.26 (p=6.32E-10) | p=4.63E-11 |
|  | FBP mcSUV<sub>R<sub>wm</sub></sub> | r=0.43 (p=1.26E-25) |  |
|  | FBP mcSUV<sub>R<sub>cb</sub></sub> and FBP mcSUV<sub>R<sub>wm</sub></sub> | r=0.79 |  |
| **AD (n=267)** |  |  |  |
| **Glucose Measure** | **Amyloid Deposition** | **Partial Correlation** | **Steigers Test** |
| **HCl** | FBP mcSUV<sub>R<sub>cb</sub></sub> | r=0.18 (p=0.0076) | p=7.95E-06 |
|  | FBP mcSUV<sub>R<sub>wm</sub></sub> | r=0.43 (p=1.85E-10) |  |
|  | FBP mcSUV<sub>R<sub>cb</sub></sub> and FBP mcSUV<sub>R<sub>wm</sub></sub> | r=0.58 |  |

**Table 3:** Partial Correlations between Longitudinal Measurements of Glucose Measures, HCI, and Amyloid Deposition: FBP mcSUVR_wm_ vs FBP mcSUVR_cb_ in CU, MCI, AD, and All Subjects (n=508) **<u>*, statistically, this test is neither valid nor needed as none of the partial correlations are significant</u>**.

|  |  | **All Subjects (n=508)** |  |
| --- | --- | --- | --- |
| **Glucose Measure** | **Amyloid Deposition** | **Partial Correlation** | **Steigers Test** |
| **HCl** | FBP mcSUV<sub>R<sub>cb</sub></sub> | r=-0.04(p=0.36) | p=2.58E-11 |
|  | FBP mcSUV<sub>R<sub>wm</sub></sub> | r=0.29(p=1.26E-11) |  |
|  | FBP mcSUV<sub>R<sub>cb</sub></sub> and FBP mcSUV<sub>R<sub>wm</sub></sub> | r=0.34 |  |
|  |  | **CU (n=166)** |  |
| **Glucose Measure** | **Amyloid Deposition** | **Partial Correlation** | **Steigers Test** |
| **HCl** | FBP mcSUV<sub>R<sub>cb</sub></sub> | r=0.10(p=0.23) | p=0.61* |
|  | FBP mcSUV<sub>R<sub>wm</sub></sub> | r=0.15(p=0.06) |  |
|  | FBP mcSUV<sub>R<sub>cb</sub></sub> and FBP mcSUV<sub>R<sub>wm</sub></sub> | r=0.19 |  |
|  |  | **MCI (n=241)** |  |
| **Glucose Measure** | **Amyloid Deposition** | **Partial Correlation** | **Steigers Test** |
| **HCl** | FBP mcSUV<sub>R<sub>cb</sub></sub> | r=-0.04(p=0.50) | p=3.94E-04 |
|  | FBP mcSUV<sub>R<sub>wm</sub></sub> | r=0.22(p<0.01) |  |
|  | FBP mcSUV<sub>R<sub>cb</sub></sub> and FBP mcSUV<sub>R<sub>wm</sub></sub> | r=0.32 |  |
|  |  | **AD (n=101)** |  |
| **Glucose Measure** | **Amyloid Deposition** | **Partial Correlation** | **Steigers Test** |
| **HCl** | FBP mcSUV<sub>R<sub>cb</sub></sub> | r=-0.15(p=0.32) | p=0.02 |
|  | FBP mcSUV<sub>R<sub>wm</sub></sub> | r=0.18(p=0.25) |  |
|  | FBP mcSUV<sub>R<sub>cb</sub></sub> and FBP mcSUV<sub>R<sub>wm</sub></sub> | r=0.70 |  |

**Figure 1:**
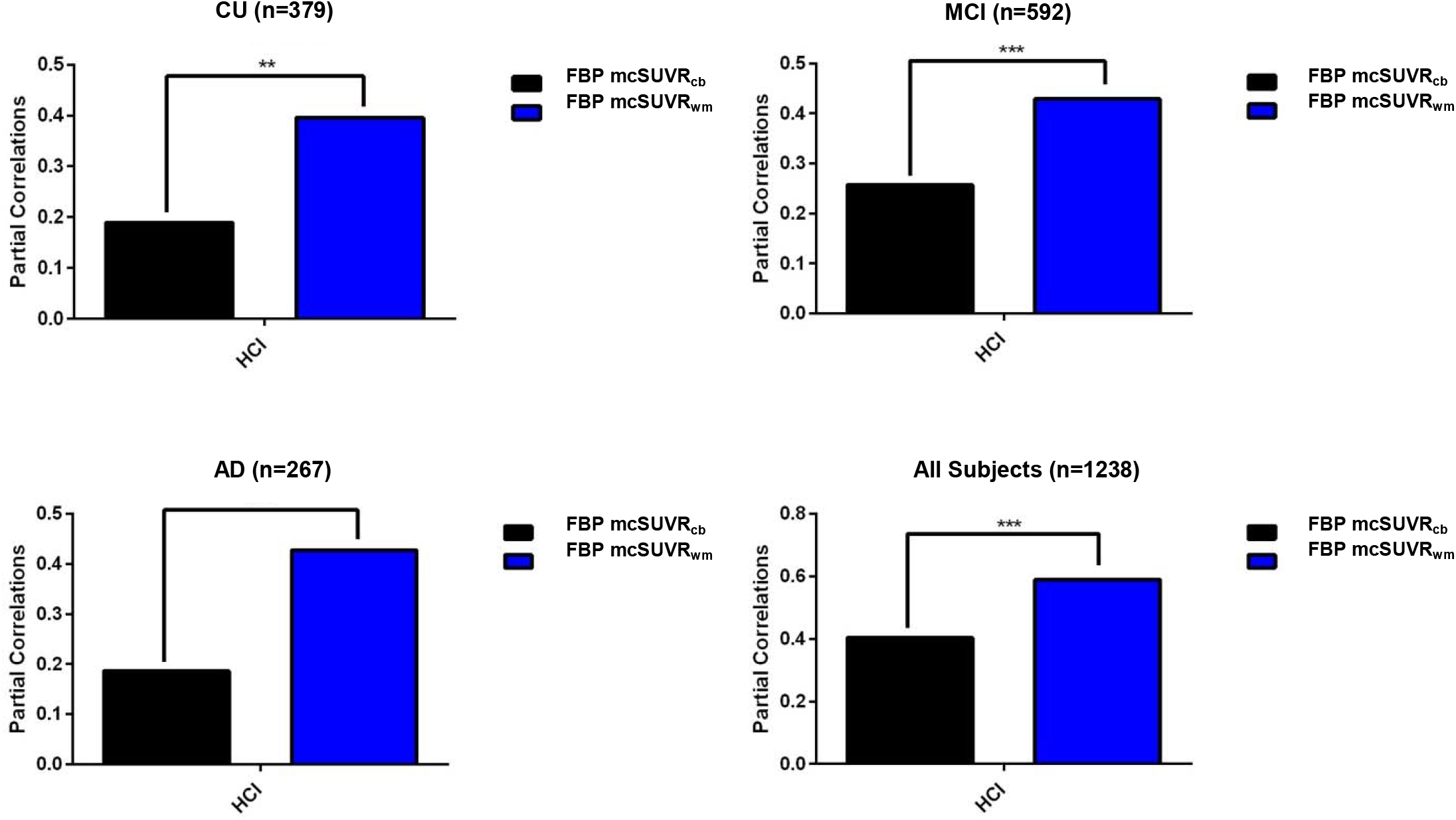
Partial Correlations between Cross-Sectional Measurements of Glucose, HCI, and Amyloid Deposition: FBP mcSUVR_wm_ vs FBP mcSUVR_cb_ in CU, MCI, AD, and All Subjects.

**Figure 2:**
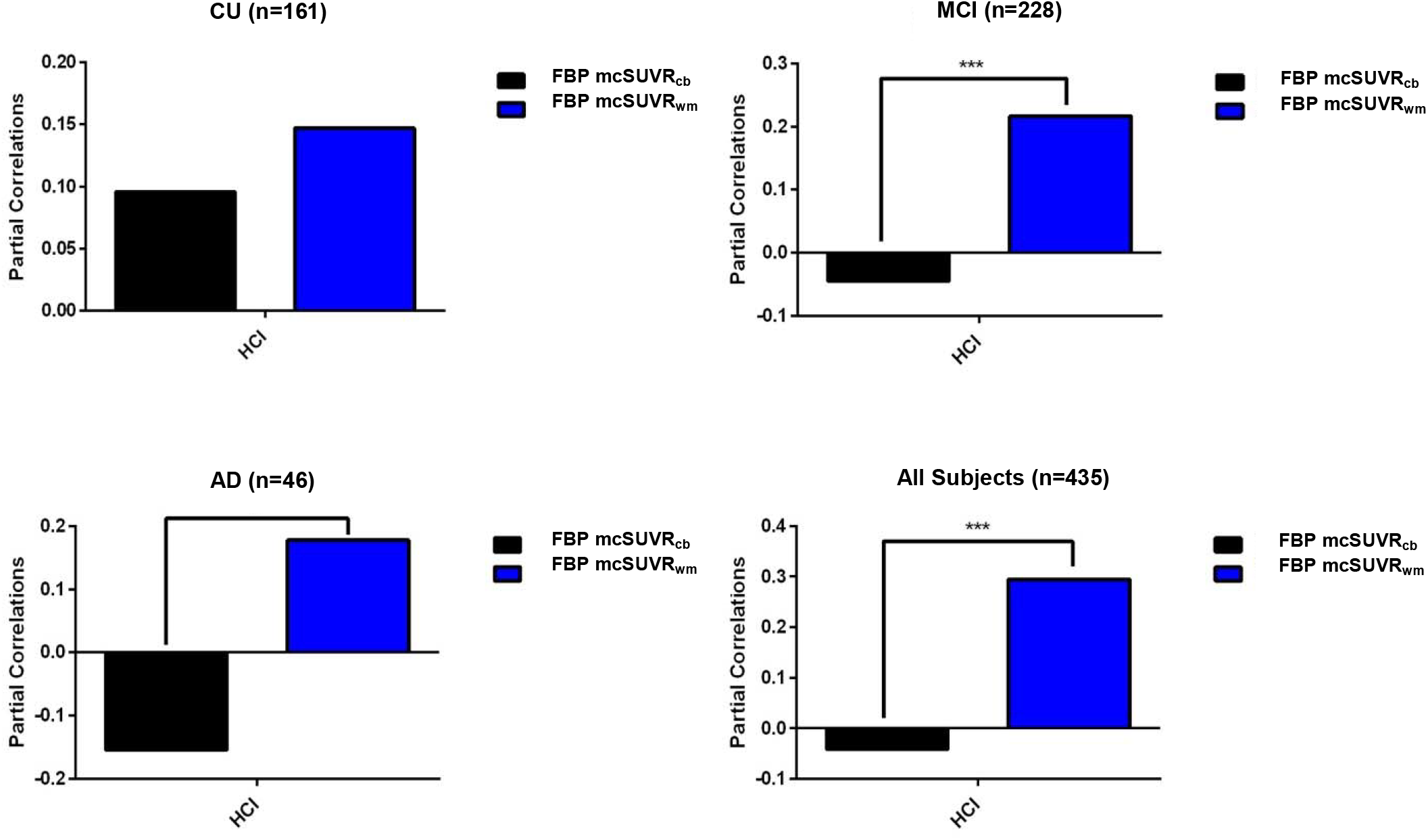
Partial Correlations between Longitudinal Measurements of Glucose Measures, HCI, and Amyloid Deposition: FBP mcSUVR_wm_ vs FBP mcSUVR_cb_ in CU, MCI, AD, and All Subjects. Of note, the negative associations noted between HCI and FBP mcSUVR_cb_ in the MCI, AD, and All Subjects were not statistically significant.

Post-hoc analysis was further conducted within AD continuum groups. Cross-sectionally, within each group, the strength of the partial correlation between HCI with FBP mcSUVR_wm_ was greater than the partial correlation between HCI and FBP mcSUVR_cb_, although not significantly in the CU group (Figure 1 and Table 2 **CU:** HCI/mcSUVR_cb_ r=0.19(p<0.001) and HCI/mcSUVR_wm_ r=0.40(p<0.001) Steiger’s Test (p<0.01); **MCI:** HCI/mcSUVR_cb_ r=0.26(p<0.001) and HCI/mcSUVR_wm_ r=0.43(p<0.001) Steiger’s Test(p<0.001); **AD:** HCI/mcSUVR_cb_ r=0.18(p<0.01) and HCI/mcSUVR_wm_ r=0.43(p=1.85E-10 Steiger’s Test(p<0.001)).

Longitudinally, within the MCI and AD cohorts, we observed stronger HCI/mcSUVR_wm_ correlations than HCI/mcSUVR_cb_ (Figure 2 and Table 3 **CU:** HCI/mcSUVR_cb_ r=0.10(p=0.23) and HCI/mcSUVR_wm_ r=0.15(p=0.06), Steiger’s Test (p=0.61); **MCI:** HCI/mcSUVR_cb_ r=-0.04(p=0.50) and HCI/mcSUVR_wm_ r=0.22(p<0.01), Steiger’s Test (p<0.001); **AD:** HCI/mcSUVR_cb_ r=-0.15(p=0.32) and HCI/mcSUVR_wm_ r=0.18(p=0.25), Steiger’s Test (p=0.02).

## Discussion

In this study, we reexamined the relationship between glucose metabolism and amyloid, using a global measure of glucose metabolism, HCI, and two different reference regions for measuring amyloid deposition (mcSUVR_wm_ and mcSUVR_cb_) for all subjects, and in CU, MCI and AD participants separately. When the cerebral white matter was used as reference region for amyloid deposition quantification, stronger associations were observed between amyloid deposition and our glucose imaging endpoint. We observed similar trends with both our cross-sectional and longitudinal analyses. In addition to the stronger associations noted here between amyloid deposition and glucose metabolism, in our companion paper, we also found stronger associations between cognition and amyloid deposition when the white matter was used as a reference region. We also note the non-significance of the HCI/FBP mcSUVR_wm_ partial correlation within the AD group. This result could be due to the small sample size (n=46) in the AD group and be explained by the plateauing of amyloid deposition at this stage.

Historically, the field believes amyloid deposition and glucose metabolism to be indirectly related, finding correlations between the two to be low and/or insignificant. Notably, these studies varied in their methods for quantifying both amyloid deposition (typically cerebellum and pons used as reference regions in SUVR calculations) and glucose metabolism. In this study, our voxel-based measure, HCI, considers the AD-specific spatial distribution pattern of glucose metabolism, while the other studies noted here examined regional or composite region-based measures of glucose metabolism. Taken together, methodologies in examining amyloid deposition and glucose metabolism may result in a difference in strength of associations, ultimately effecting our biological understanding of AD pathology.

Amyloid beta quantification depends on accurate selection of (1) reference regions and (2) regions of interest to ultimately derive SUVRs. Reference regions are selected on the assumption that the region is devoid of amyloid binding, has similar tissue characteristics as the region of interest, and has low variability across diagnostic groups. Although the cerebellum has been cross validated using a plasma input approach and is relatively free of amyloid deposition (with the exception of rare familial forms of AD), technical limitations exist with this reference region. For example, accurate segmentation of this reference region can be influenced by truncation of field of view in the lower part of the brain plus lower sensitivity due to this field of view (*8,9*). Cerebellar regions are more susceptible to noise from adjacent tissue with specific amyloid tracer binding (*8*). On the other hand, the cerebral white matter is a larger region with the potential to reflect non-specific signal intensity more adequately. Cerebral white matter is also located more central in the field of view, thus less susceptible to noise and artifacts (*8*). In addition, our use of this cerebral white matter reference region further away from cortical areas, limits partial volume effects or spillover of cortical amyloid signal (*8*).

It has been noted that some cortical amyloid signal may be included as part of subcortical white matter (*10*). However, our white matter reference region avoided selecting white matter regions close to grey matter, thereby limiting signal contamination from the cortical regions. Blood flow in WM has further been reported to be slower than grey matter and slower clearance of tracers in WM have been reported in molecular studies(*11*). Additionally, although Lowe et al. found that WM PiB tracer uptake increases with age and varies with baseline gray matter amyloid beta deposition, they noted (i) a lower level of annual tracer uptake change in eroded subcortical WM ROIs was observed in both longitudinal and cross-sectional studies and (ii) that such minimal increase made it is suitable to use the white matter reference region for short follow-up studies (*10*). Furthermore, a study by Landau et al. showed that the use of subcortical white matter as a reference region could detect amyloid beta changes that were more physiologically plausible and more likely to increase over time compared to the cerebellum or pons alone (*2*). Wang et al. in 2021, further found that white matter reference region, when compared to the whole cerebellar reference region, was better able to identify diagnostic groups of SUVRs (*8*). All three of these studies provide supportive arguments lessening the concern of using eroded cerebral white matter reference region in our results(*2,8,10,12*). Moreover, the use of cerebral reference ROIs in the same axial field of view could generate SUVRs with reduced variability due to decreased scatter and improve data quality to track an increase in longitudinal fibrillar amyloid beta plaque (*12*). Importantly, reference region selection also effects AD clinical trials. Chiao et al. in 2019 showed use of subcortical white matter as a reference region generated larger effect sizes than the use of whole cerebellum as a reference region in the Phase 1b Prime Study of Aducanamab (*13*).

We acknowledge the existence of other possible factors contributing to the greater association observed between HCI and mcSUVR using white matter as a reference region. In addition to its use for detection of amyloid, amyloid PET tracers are currently being used as measures of white matter changes, given the structural similarities between amyloid beta and myelin basic protein (*14*). Some studies have shown a decline in amyloid PET tracer uptake in white matter with AD disease progression (*14,15*). This is thought to reflect changes in white matter integrity/fiber necrosis, vascular damage, edema, or inflammation (*14-16*). Other studies have shown amyloid PET tracer uptake in white matter declines with age (*10*). With these findings, our results should be interpreted with caution. On the other hand, our white matter reference region is a subset of whole white matter. **This specific subset of white matter has shown stability over time and across disease severity (Villemagne et al. 2016 abstract submission, Human Amyloid Imaging Conference**). Additionally, including white matter in the reference region was found to counteract the effects of white matter signal spill into cortical target regions in amyloid PET tracers (*17*).

Biologically, reductions in glucose metabolism can be seen several decades before AD pathology begins(*18*). In cognitively unimpaired APOE4 carriers, Reiman et al. noted abnormally low glucose cerebral metabolic rates in the posterior cingulate, parietal, temporal, and prefrontal cortex (*18*). In the posterior cingulate, each of the six layers of this region showed a decline in cytochrome oxidase activity, a measure of oxidative measure capacity in mitochondria in AD patients when compared to nondemented controls (*19*). Our results are supported by these previous studies. At baseline, cross-sectionally, CU, MCI, and AD individuals show similar strengths of associations between glucose metabolism and amyloid deposition (when white matter is used as a reference region). Longitudinally, significant associations between glucose metabolism and amyloid deposition were only noted in the MCI group and not in the CU group (majority amyloid negative individuals) and AD groups. This could indicate that in the “active” declining state of MCI, when amyloid deposition is reaching its peak, metabolic glucose decline is related to AD pathological changes or amyloid in our study. Whether glucose metabolism changes are related to amyloid/tau individually or a combination of these and other pathophysiological changes remains unknown. However, we continue to postulate that the changes in glucose metabolism reflect a reduction in density and/or activity of terminal neuronal fields or peri-synaptic astroglial cells that project to the brain regions more susceptible to metabolic reductions in AD.

## Limitations

One limitation of our study was we only investigated the relationship between amyloid deposition and glucose metabolism without inclusion of other relevant pathophysiologic phenomenon such as tau deposition. Tau deposition has been found to be more closely associated with glucose metabolism in multiple studies (*20,21*). Therefore, a follow-up study is planned to compare the strengths of the relationships between tau deposition and glucose metabolism compared to amyloid deposition using white matter as the reference region for SUVR quantification. Furthermore, we only examined FBP-measured amyloid deposition and have not examined the role of other amyloid PET tracers such as Pittsburgh Compound B (PiB) or other 18-F tracers. As discussed above, PiB PET tracer signal in white matter increases with age and can vary based on amyloid beta deposition in gray matter (even in eroded white matter regions) (*10*) therefore PiB specifically could underestimate cerebral amyloid deposition.

Importantly, our study is a correlational analysis therefore, we cannot draw any conclusions on causality. Altogether, our results suggest a closer relationship between amyloid deposition and glucose metabolism than previously observed. This study provides further support for the use of FBP mcSUVR_wm_ measurements in the detection and tracking of AD.

## Data Availability

All data produced in this work are contained in the manuscript.

https://adni.loni.usc.edu/

